# An infectious disease model with asymptomatic transmission and waning immunity

**DOI:** 10.1101/2023.10.24.23297464

**Authors:** Sophia Y. Rong, Alice X. Li, Shasha Gao, Chunmei Wang

## Abstract

Infectious diseases present persistent challenges to global public health, demanding a comprehensive understanding of their dynamics to develop effective prevention and control strategies. The presence of asymptomatic carriers, individuals capable of transmitting pathogens without displaying symptoms, challenges conventional containment approaches focused on symptomatic cases. Waning immunity, the decline in protective response following natural recovery or vaccination, introduces further complexity to disease dynamics. In this paper, we developed a mathematical model to investigate the interplay between these factors, aiming to inform strategies for the management of infectious diseases. We derived the basic reproduction number for the model and showed that the disease would die out when this number falls below 1. We obtained a formula to estimate the relative contributions of asymptomatic and symptomatic transmission to the basic reproduction number, which remains unchanged when vaccination is included in the model. Through computer simulations with parameter values tailored for COVID-19 and sensitivity analysis, we demonstrated that population susceptibility significantly impacts the timing and magnitude of infection peaks. Populations with lower susceptibility experience delayed and less severe outbreaks. Vaccination was shown to play a crucial role in disease control, with an increased vaccination rate, extended immunity, and heightened vaccine efficacy proving pivotal. However, the effectiveness of these strategies hinges on maintaining a low vaccine escape proportion. Taken together, this study underscores the need for multifaceted, adaptable approaches to infectious disease management, highlighting the central role of vaccination in mitigating disease spread. Further research and validation with disease-specific data will enhance parameter estimates, improve model predictions, and inform evidence-based disease control strategies.

## 1. Introduction

Infectious diseases continue to pose significant challenges to global public health, necessitating concerted efforts from researchers and healthcare practitioners to develop comprehensive strategies for understanding, preventing, and managing them [5, 25]. Two specific features of infectious diseases have recently garnered considerable attention due to their complexity and potential implications: asymptomatic transmission [40] and waning immunity [10]. These phenomena have the capacity to exert profound influences on disease dissemination patterns, modulate the efficacy of interventions, and add layers of complexity to public health response efforts.

Traditionally, the identification and containment of infectious diseases have revolved around symptomatic cases — individuals who exhibit discernible clinical signs of infection. However, the presence of asymptomatic carriers, individuals who harbor and transmit the pathogen without displaying overt symptoms, has challenged this conventional approach. The COVID-19 pandemic vividly demonstrated the substantial impact of asymptomatic carriers in propagating the causative agent, SARS-CoV-2 [18, 33]. Other infectious diseases also exhibit asymptomatic transmission, including influenza [24], HIV [36], hepatitis B and C [22], tuberculosis [15], measles [3], herpes simplex virus [16], malaria [6], among others. Asymptomatic transmission may stand as a formidable contributor to disease propagation, allowing the pathogen to silently infiltrate and establish itself within communities. Such stealthy transmission often evades detection, rendering conventional control measures less effective and making it crucial to develop strategies that account for this hidden reservoir of infection.

Immunity acquired following an infection or vaccination is not always an enduring shield against reinfection. Over time, the protective response generated by the immune system can wane, leaving individuals susceptible to subsequent infections. The phenomenon of waning immunity is observed in various infectious diseases, introducing an additional layer of complexity into the dynamics of infectious diseases [4, 12, 19, 37, 44, 47]. It can lead to scenarios where a population’s overall immunity decreases, potentially giving rise to resurgences of the disease even after it appeared to be under control. The interplay between waning immunity and the introduction of asymptomatic carriers can further amplify the intricacies of disease dynamics and pose challenges for maintaining long-term population-level immunity [11, 9, 27, 32].

Understanding the interactions between asymptomatic transmission and waning immunity is pivotal for comprehending the overall trajectory of infectious diseases. These features can interact in intricate ways, affecting the overall disease burden, the potential for recurrent outbreaks, and the effectiveness of vaccination and control strategies [43]. Mathematical modeling has emerged as a powerful tool to unravel the intricate dynamics of infectious diseases. By constructing mathematical frameworks that capture the interplay between hosts, pathogens, and environmental factors, researchers gain insights into disease transmission patterns, assess intervention strategies, and predict potential outcomes. For example, Moghadas et al. [30] and He et al. [23] found that asymptomatic transmission of COVID-19 could substantially contribute to the spread of the virus and that containment measures should consider this silent transmission. Althouse and Scarpino [2] explored the role of asymptomatic carriers in the resurgence of pertussis (whooping cough), highlighting that even if a high proportion of individuals are vaccinated, asymptomatic transmission can still sustain the disease in the population. Models have also been used to study the roles of asymptomatic transmission for other diseases, such as Zika infection [31], measles [32, 46], respiratory syncytial virus (RSV) [35, 42], etc.

Mathematical models have also been developed to study waning immunity. Mossong et al. [32] demonstrated that a single-dose routine vaccination strategy would not be sufficient for the elimination of the measles virus, provided the basic reproduction number among vaccinated individuals surpasses a certain threshold. This underscores the importance of determining both the intensity and duration of infectiousness in vaccinated individuals. López et al. [28] analyzed more than 20 years of dengue virus infection epidemics in the isolated French Polynesian islands and found that lifelong serotype-specific immunity may not occur and that including waning immunity in models improved their accuracy in capturing epidemic dynamics. Woolthuis et al. [45] showed that variation in the duration of immunity can shape influenza epidemics and impact the long-term effectiveness of vaccination. These models provide a unique lens through which to investigate complex scenarios that are often challenging to dissect solely through empirical studies.

In this paper, we develop and analyze an infectious disease model, aiming to shed light on the complex interplay between asymptomatic transmission and waning immunity, among other factors. Our motivation comes from the pressing need to address real-world scenarios where asymptomatic individuals may play a significant role in transmission dynamics, and where immunity wanes over time, potentially leading to resurgences of infection. Through our exploration, we aim to contribute to the understanding of these multifaceted dynamics and provide insights that can inform decision-making and policy development for disease control and prevention.

This paper is structured as follows: section 2 outlines the model formulation, defining the compartments and parameters governing the dynamics of infectious diseases with asymptomatic transmission and waning immunity. In section 3, we perform a detailed analysis of the model, including the derivation of the basic reproduction number, study the model’s steady states and their stability, and evaluate the relative contribution from the asymptomatic and symptomatic transmissions. In section 4, we present the results of numerical investigations, employing COVID-19 as a case study to calibrate parameter values and illustrate disease spread dynamics under various scenarios. We explore how various factors, such as asymptomatic transmission, waning immunity, and vaccination effectiveness, can affect the disease dynamics. The conclusions follow in section 5.

## 2. Model development

We divide the total human population into six compartments: susceptible (*S*), vaccinated (*C*), exposed (*E*), asymptomatic infected (*A*), symptomatic infected (*I*), and recovered (*R*). The model is described by the following system of ordinary differential equations,

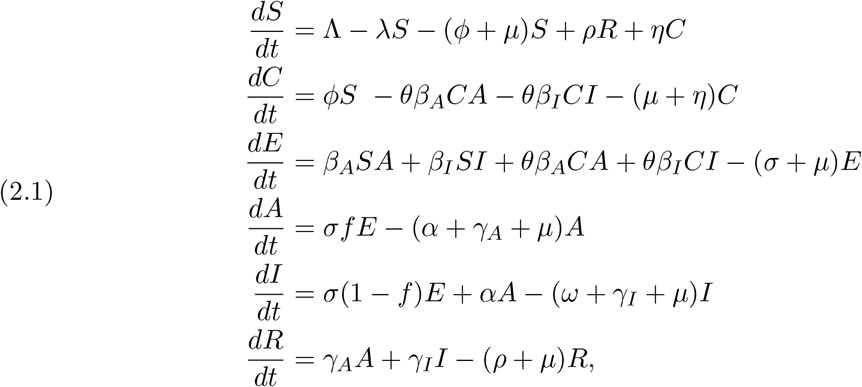

with *λ* = *β*_*A*_*A* + *β*_*I*_*I* representing the force of infection. In the model, the parameter Λ represents the generation rate of the susceptible population, and *μ* denotes the natural death rate. Susceptible individuals can become infected either by asymptomatic infected individuals at a rate of *β*_*A*_*SA* or by symptomatic infected individuals at a rate of *β*_*I*_*SI*, thereby entering the exposed stage, during which they are not infectious. Susceptible individuals are assumed to be vaccinated at a per capita rate of *ϕ*. Because the vaccine may not be perfect, individuals can still become infected following vaccination, with the extent of protection represented by the vaccine escape proportion denoted as *θ ∈* (0, 1). When *θ* = 0, the vaccine offers perfect protection, and no vaccinated individuals will be infected. In contrast, when *θ* = 1, the vaccine provides no protection, and vaccinated individuals have the same infection rate as fully susceptible individuals. Vaccinated individuals can revert to a susceptible state at a rate of *η* due to vaccine’s *waning immunity*.

Exposed individuals are assumed to progress to the next stage at a rate of *σ*, becoming either asymptomatic with a fraction *f* or symptomatic with a fraction of 1−*f*. Asymptomatic infected individuals may either become symptomatic at a rate of *α* or recover at a rate of *γ*_*A*_. Symptomatic infected individuals have a disease-induced mortality rate of *ω* and a recovery rate of *γ*_*I*_. Naturally recovered individuals can also revert to a susceptible state at a rate of *ρ* due to *waning immunity*. For simplicity of notation, in the model, we combine parameters and denote:

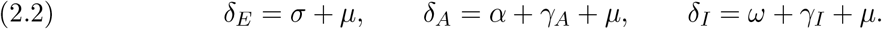

A diagram of model (2.1) is presented in Figure 1. The descriptions of variables and parameters are summarized in Table 1.

**Table 1.** Model variables and parameters.

| Symbol | Meaning | Baseline value | Unit | Source |
| --- | --- | --- | --- | --- |
| **Variables** |  |  |  |  |
| $S(t)$ | Susceptible population | | | |
| $C(t)$ | Vaccinated population | | | |
| $E(t)$ | Exposed population | | | |
| $A(t)$ | Asymptomatic infected population | | | |
| $I(t)$ | Symptomatic infected population | | | |
| $R(t)$ | Recovered population | | | |
| $N(t)$ | Total population | | | |
| **Parameters** |  |  |  |  |
| $\Lambda$ | Recruitment rate of susceptible | 11528 | person/day | [21] |
| $\mu$ | Natural death rate | $1/(79 \times 365)$ | 1/day | [21] |
| $\beta_A$ | Asymptomatic transmission rate | $1.311 \times 10^{-9}$ | 1/(person × day) | [21] |
| $\beta_I$ | Symptomatic transmission rate | $4.334 \times 10^{-9}$ | 1/(person × day) | [21] |
| $\sigma$ | Transition rate from $E$ to $A$ or $I$ | 1/5.44 | 1/day | [48] |
| $\rho$ | Waning rate of natural immunity | 1/238 | 1/day | [48] |
| $f$ | Proportion of asymptomatic infection | 35% | unitless | [34] |
| $\alpha$ | Transition rate from $A$ to $I$ | 0.33 | 1/day | [21] |
| $\gamma_A$ | Recovery rate of individuals in class $A$ | 0.094 | 1/day | [21] |
| $\gamma_I$ | Recovery rate of individuals in class $I$ | 0.094 | 1/day | [21] |
| $\omega$ | Disease-induced death rate | 0.017 | 1/day | [21] |
| $\phi$ | Vaccination rate | 1/30 | 1/day | Estimated |
| $\eta$ | Waning rate of vaccine-induced immunity | 1/127 | 1/day | [48] |
| $\theta$ | Vaccine escape proportion | 0.3 | unitless | [48] |

**Figure 1.**
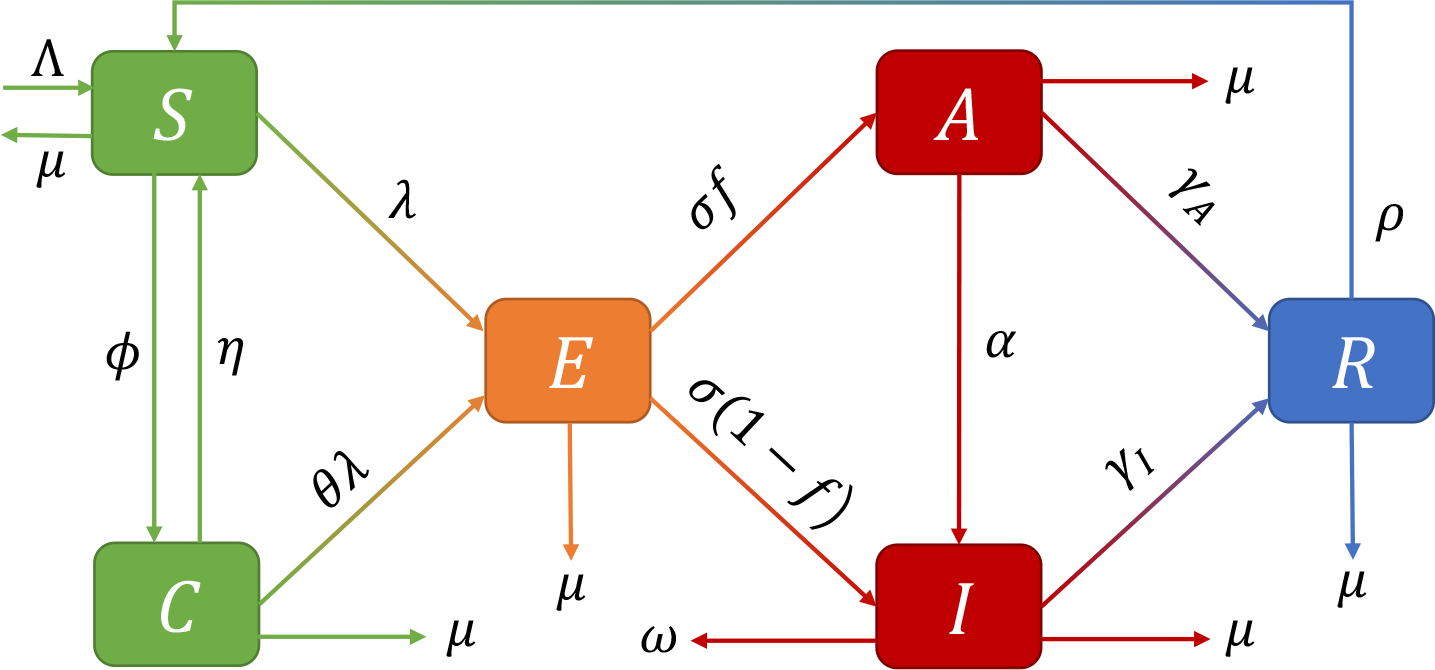
Flow diagram of model (2.1). The population is divided into 6 compartments: susceptible individuals (S), vaccinated individuals (C), exposed individuals (E), asymptomatic infected individuals (A), symptomatic infected individuals (I), and recovered individuals (R). Detailed model parameter descriptions are given in Table 1.

From model (2.1), we can observe that all the derivatives evaluated at the state 0 satisfy

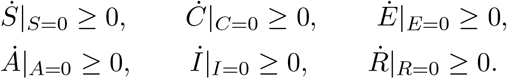

Hence, if the initial conditions remain within the closed positive hyperspace 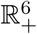, the solution will also remain in 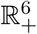 for the entire duration, i.e., 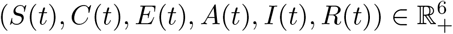 for any time *t*. Defining *N*(*t*) = *S*(*t*) + *C*(*t*) + *E*(*t*) + *A*(*t*) + *I*(*t*) + *R*(*t*) as the total population, and summing the equations in (2.1), we obtain

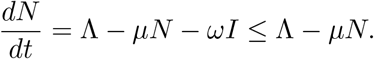

It follows that

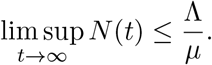

Hence, we define the domain of the model (2.1) as

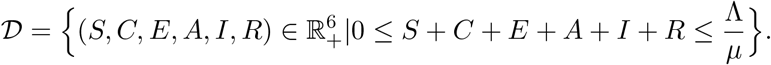

𝒟 is positively invariant for system (2.1). In other words, trajectories with initial conditions in 𝒟 will remain within 𝒟 for all time. Using a method similar to that in [20], we can show the existence of a unique solution of system (2.1) in 𝒟 for any initial conditions in 𝒟. Thus, system (2.1) is both epidemiologically and mathematically well-posed.

## 3. Model analysis

### 3.1 Model without vaccination

We begin the analysis of the model without vaccination, which can be simplified as follows

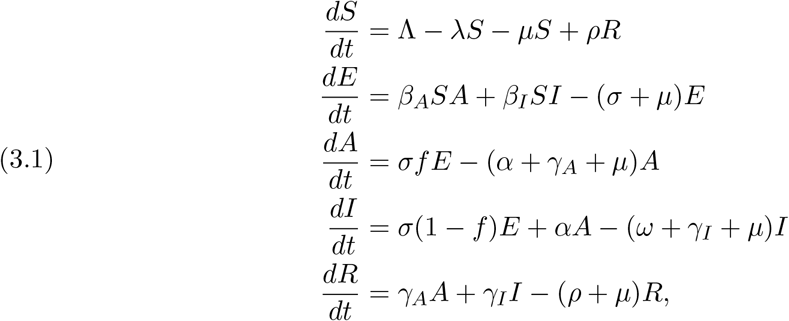

with *λ* = *β*_*A*_*A* + *β*_*I*_*I* representing the force of infection.

#### 3.1.1. Basis reproduction number

The basic reproduction number, widely used in epidemiology, represents the average number of secondary infections produced by one infected individual in a completely susceptible population. We derive the basic reproduction number using the next generation approach as follows [41]. Only considering the infected classes *x* = (*E, A, I*) and using the notations in (2.2), we get the system 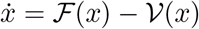, where

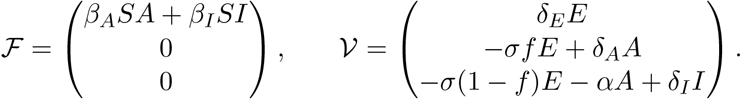

Evaluating the Jacobian matrices of ℱ and 𝒱 at the disease-free equilibrium for model (3.1), i.e. ℰ^0^ = (*S*^0^, 0, 0, 0, 0) where *S*^0^ is equal to Λ/*μ*, we get

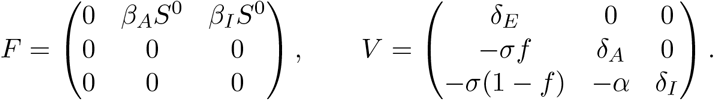

The inverse of matrix *V* is

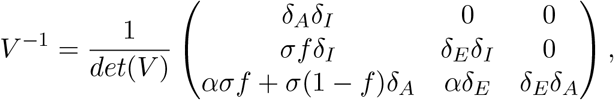

where the matrix determinant is *det*(*V*) = *δ*_*E*_*δ*_*A*_*δ*_*I*_. It follows that

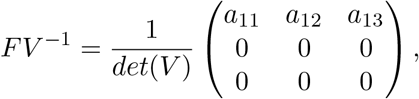

where

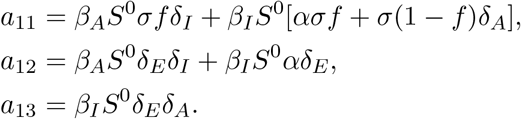

According to [41], the basic reproduction number ℛ_0_ is the spectral radius of the matrix *FV* ^−1^. Therefore,

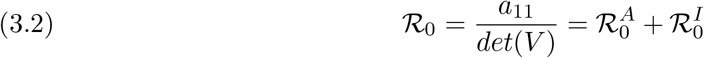

with

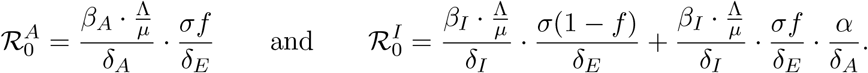

In the term 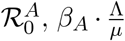 represents the rate at which one asymptomatic infected individual generates exposed individuals in an entirely susceptible environment per unit of time, 1/*δ*_*A*_ is the average duration an individual spends in the asymptomatic infected group *A*, and *σf*/*δ*_*E*_ represents the fraction of infected individuals who transition from the exposed group *E* to the asymptomatic infected class *A*. Therefore, 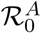 quantifies the number of secondary asymptomatic infected individuals produced by one infectious individual while in group *A*.

Similarly, the term 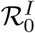 represents the number of secondary symptomatic infected individuals generated by one infectious individual while in class *I*. The two terms in this expression account for the transition of infected individuals from class *E* either directly to *I* or via *A* before entering *I*. Taken together, the expression for the basic reproduction number ℛ_0_ highlights the importance of reducing both asymptomatic and symptomatic infected individuals.We can further evaluate the relative contribution to the basic reproduction number from asymptomatic and symptomatic transmissions. This ratio is given by

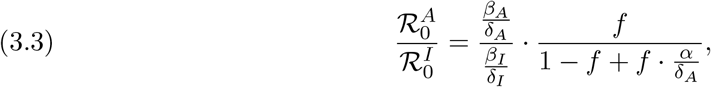

which depends on the transmission rates (*β*_*A*_ and *β*_*I*_), proportion of asymptomatic infection (*f*), transition rate (*α*) from asymptomatic to symptomatic, and removal rates from compartments (*δ*_*A*_ and *δ*_*I*_).

#### 3.1.2. Existence and stability of equilibria

We calculate the equilibrium point of the model (3.1). Let all the derivatives in the system be 0. From the third, fourth, and fifth equation of model (3.1), we get the equilibrium in terms of *A*

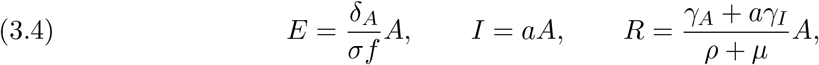

where

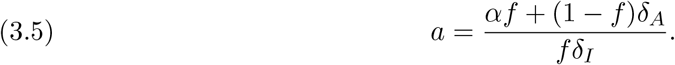

Using the above expression for *a* and performing a straightforward calculation, we can rewrite the basic reproduction number (3.2) as

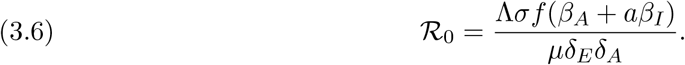

Substituting (3.4) into the second equation of (3.1), we have

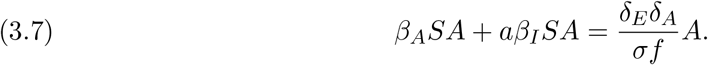

There are two cases. The first is when *A* = 0. It follows that *S* = Λ/*μ, E* = 0, *I* = 0, and *R* = 0. Thus, we have the disease-free equilibrium point for model (3.1), which is denoted by ℰ^0^ and given by

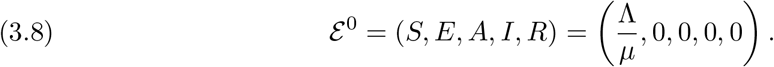

When *A ≠* 0, dividing Eq. (3.7) by *A* leads to

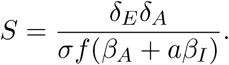

Using the basic reproduction number (3.6), the above expression for *S* can be rewritten as

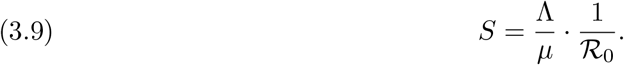

Let *N* = *S* + *E* + *A* + *I* + *R* be the total population. Adding all the equations in (3.1) yields

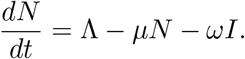

At the equilibrium, the above derivative should be zero, leading to

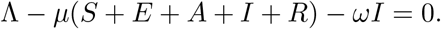

Using (3.4), we obtain

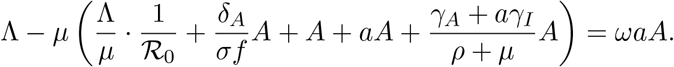

Solving for *A*, we have

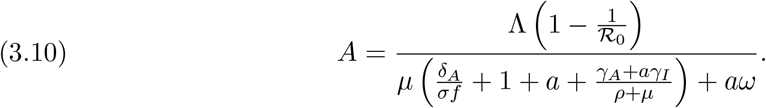

It follows from (3.4), (3.9) and (3.10) that all the components of the positive equilibrium or endemic equilibrium (*S, E, A, I, R*) are in the interval (0, Λ/*μ*) if and only if ℛ_0_ *>* 1. We summarize the above results in the following theorem.

##### Theorem 3.1

*Model*(3.1) *always admits a unique disease-free equilibrium ℰ* ^0^, *given by*(3.8). *The endemic equilibrium ℰ*^*\**^ = (*S*^*\**^, *E*^*\**^, *A*^*\**^, *I*^*\**^, *R*^*\**^) *exists if and only if* ℛ*R*_0_ *>* 1, *and it is given by*

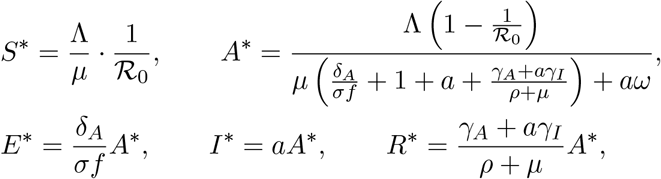

*where* ℛ_0_ *and a are given in*(3.2) *and*(3.5), *respectively*.

We are interested in the scenario where the disease dies out, i.e., when the system converges to the disease-free equilibrium point ℰ^0^. We have the following result.

##### Theorem 3.2

*When* ℛ_0_ < 1, *the disease-free equilibrium ℰ*^0^ *is locally asymptotically stable*.*When* ℛ_0_ *>* 1, ℰ^0^ *is unstable*.

*Proof*. By evaluating the Jacobian matrix of system (3.1) at the disease-free equilibrium ℰ^0^ and denoting the eigenvalue as *ζ*, we get the characteristic equation, given by

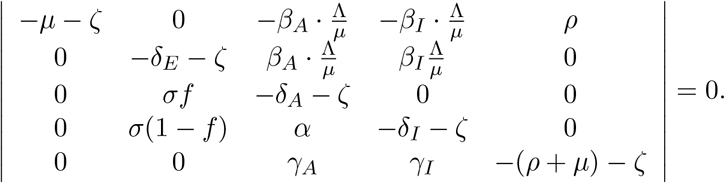

There are two negative eigenvalues, *ζ* = −*μ* and *ζ* = −(*ρ* + *μ*). The remaining eigenvalues are determined by

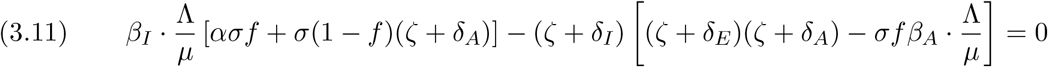

When ℛ_0_ *>* 1, we denote the left side of (3.11) as *F*(*ζ*), which is a continuous function on [0, +*∞*). On the one hand,

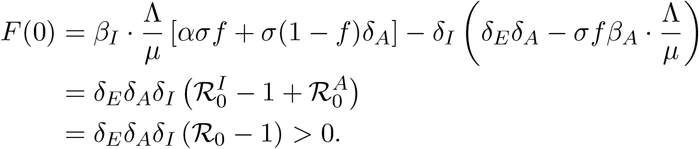

On the other hand, *F*(*ζ*) *→* −*∞* as *ζ →* +*∞*. According to the intermediate value theorem, *F*(*ζ*) has at least one positive root, which indicates that the disease-free equilibrium *ℰ*^0^ is unstable.

When ℛ_0_ < 1, we rewrite (3.11) as

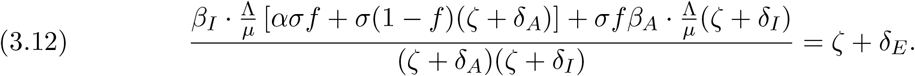

We denote the left and right side of (3.12) as *G*(*ζ*) and *H*(*ζ*), respectively. Suppose the real part of the eigenvalue is *Re*(*ζ*) ≥ 0. We evaluate the modulus of the left side and obtain

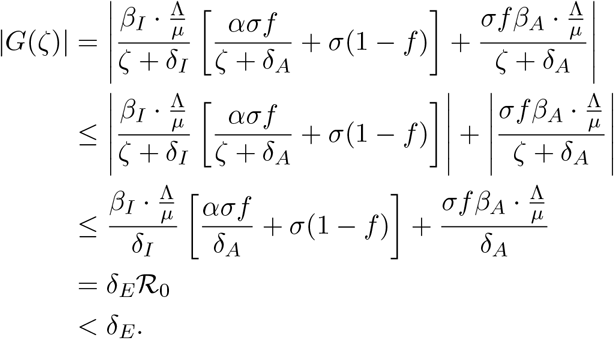

However, the modulus of the right side is |*H*(*ζ*)| ≥ *δ*_*E*_ when *Re*(*ζ*) ≥ 0. This results in a contradiction in (3.12). Thus, all the roots of the characteristic equation (3.12) have negative real parts and the disease-free equilibrium *ℰ*^0^ is locally asymptotically stable. This completes the proof.

**Remark**. By evaluating the Jacobian matrix at the endemic equilibrium point *ℰ*^*\**^ = (*S*^*\**^, *E*^*\**^, *A*^*\**^, *I*^*\**^, *R*^*\**^) as provided in Theorem 3.1, we can derive the characteristic equation. After extensive computation, this equation is expressed as follows:

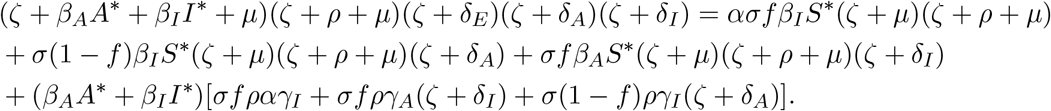

While we can expand the products in the above equation and derive some general stability conditions for the endemic equilibrium using the Routh-Hurwitz Criterion, expressing these conditions succinctly in terms of the basic reproduction number ℛ_0_ is challenging, if not impossible. Therefore, we will not present the general stability conditions for the endemic equilibrium *ℰ*^*\**^.

### 3.2. Model with vaccination

In this section, we study the model with vaccination (2.1). Similar to model (3.1), system (2.1) always has a disease-free equilibrium

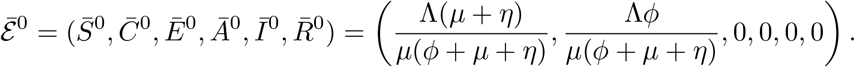

Applying a similar method, we can derive the basic reproduction number for model (2.1) with vaccination. Because it is under the influence of vaccination measures, we refer to it as the control reproduction number, which is given by

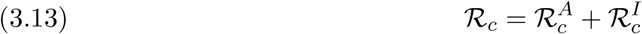

where

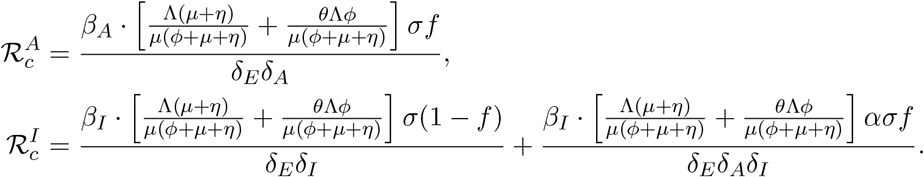

The control reproduction number ℛ_*c*_ shares a similar biological interpretation with the basic reproduction number ℛ_0_. 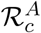 represents the contribution from asymptomatic transmission, whereas 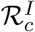 represents the contribution from symptomatic transmission. It’s worth noting that when the vaccination rate *ϕ* = 0, the control reproduction number is precisely the same as the basic reproduction number stated in (3.2).

We also evaluate the relative contribution to the control reproduction number from asymptomatic and symptomatic transmissions, and obtain

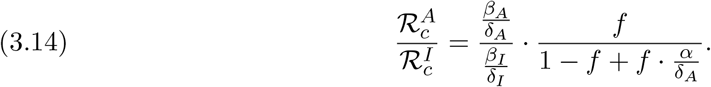

This is consistent with the ratio in (3.3), indicating that the inclusion of vaccination does not alter the relative contributions of asymptomatic and symptomatic transmissions.

Due to the complexity of model (2.1), we will conduct numerical investigations to explore the existence and stability of its equilibria, as well as the influence of various parameters on the disease dynamics.

## 4. Numerical investigations

In this section, we use COVID-19 as an example to calibrate parameter values, illustrate the dynamics of disease spread in various scenarios, and evaluate how parameters — particularly the vaccination rate, waning immunity, asymptomatic transmission, and the vaccine escape proportion — affect the spread of the disease.

Given that the life expectancy in the US population is approximately 79 years [7], we set the natural death rate to *μ* = 1/(79 × 365) per day. The total US population in 2021 is approximately *N*_0_ = 332, 398, 949, and it is assumed to remain relatively stable during the period of the numerical simulation. Consequently, the rate of birth is approximately the rate of death, which allows us to get an estimation of the recruitment rate Λ = *N*_0_*μ*, measured in individuals per day. The other parameter values are listed in Table 1, with most of them chosen based on data fitting a model to COVID-19 cases [21].

Using the parameter values from Table 1, we numerically solved model (2.1) and plotted the predicted dynamics of asymptomatic and symptomatic infected individuals in Figure 2(a) and (b) respectively. In each figure, two scenarios were compared: one assuming a high initial proportion of susceptible individuals (80%) and the other assuming a low initial susceptibility (5%). It was observed that, in both scenarios, both asymptomatic and symptomatic infected individuals converged to a steady state after reaching the peak (solid lines in Figure 2). The calculation of the control reproduction number using the formula (3.13) yielded ℛ_*c*_ = 5.352, which supports the hypothesis that the disease becomes endemic (i.e. the endemic equilibrium of the model is asymptotically stable) when ℛ_*c*_ exceeds 1. Although the initial conditions (80% vs. 5% susceptibility) did not affect the final endemic state, the results highlighted the significant impact of population susceptibility on the model’s dynamics, with populations exhibiting lower susceptibility experiencing lower and delayed infection peaks.

**Figure 2.**
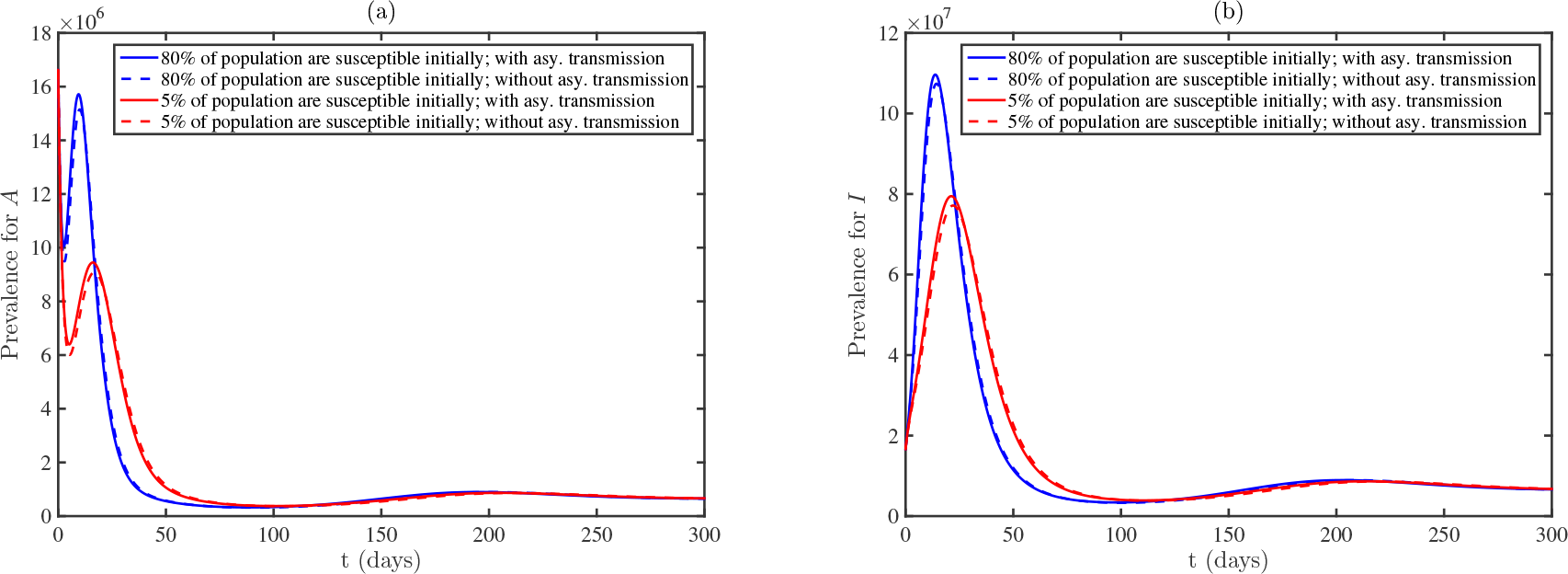
Convergence to the endemic equilibrium for model (2.1) with ℛ_c_ = 5.352. The parameter values are detailed in Table 1. The solid lines depict the disease dynamics with varying proportions of susceptible individuals at the outset. The dashed lines represent the scenario without asymptomatic transmission.

We further evaluated the contribution of asymptomatic transmission to the disease dynamics. For comparison, we plotted the predicted dynamics assuming no asymptomatic transmission (dashed lines in Figure 2), i.e. *β*_*A*_ = 0, while keeping other parameter values unchanged. It was found that there was no significant difference between the predictions with and without asymptomatic transmission. This finding was consistent with the calculation of the relative contribution using the formula (3.14). Using the parameter values listed in Table 1, the relative contribution from asymptomatic and symptomatic transmissions, given by the ratio 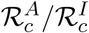, was approximately 3%. Even when we set *β*_*A*_ to be equal to *β*_*I*_, the ratio remained at approximately 10%. These results suggest that asymptomatic transmission does not contribute significantly to the total transmission.

The above scenario shows the persistence of the disease when the control reproduction number ℛ_*c*_ exceeds 1. If we increase the vaccination rate, decrease the waning rate of vaccine-induced immunity, and reduce the vaccine escape proportion each by a factor of 10, i.e., *ϕ* = 1/3, *η* = 1/1, 270, *θ* = 0.03, while keeping all other parameter values unchanged as listed in Table 1, then the control reproduction number is reduced to ℛ_*c*_ = 0.399. Figure 3 illustrates the convergence to the disease-free equilibrium for model (2.1) in this case. Panels (a) and (b) depict the prevalence of individuals in compartments *A* and *I*, respectively. As in Figure 2, we considered different initial conditions for susceptible individuals. The results indicate that the disease dies out in both scenarios when ℛ_*c*_ < 1. The model also predicts a lower peak in the prevalence of symptomatic infected individuals whenever a smaller proportion of individuals are assumed to be susceptible at the outset.

**Figure 3.**
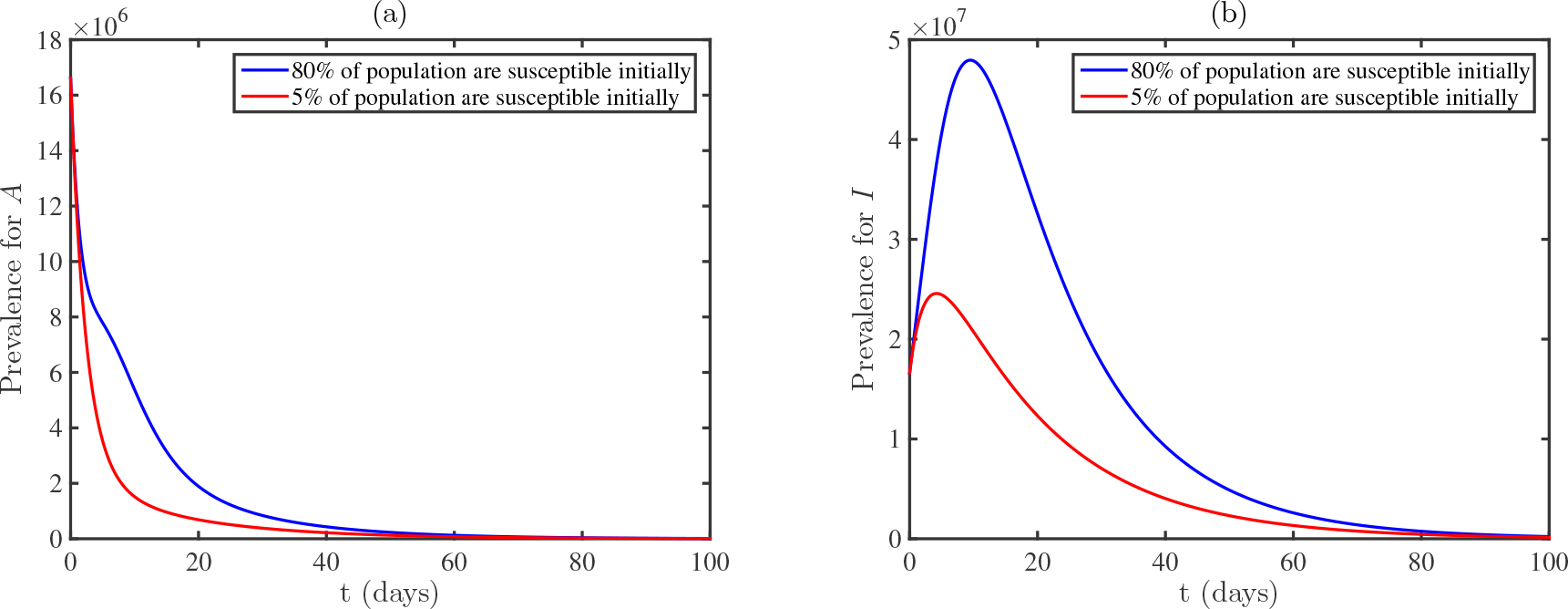
Convergence to the disease-free equilibrium for model (2.1) when ℛ_c_ = 0.399. Panels (a) and (b) display the prevalence for compartments A and I respectively. We set ϕ = 1/3, η = 1/1270, and θ = 0.03, while all other parameter values are provided in Table 1.

To understand how the changes in model parameters affect the spread of the disease, we calculated the Partial Rank Correlation Coefficients (PRCCs), which can identify which model input parameters have a strong influence on the output variable of interest. Figure 4 displays the PRCCs for the control reproduction number *R*_*c*_ (left panel) and the combined level of active infections *A* + *I* on the 25th day (right panel), with respect to all model parameters. We chose day 25 because the disease peak occurs approximately at that time. The baseline values for these parameters are listed in Table 1. Most parameter ranges in the sensitivity test extend by 20% in both directions from their respective baseline values. The results strongly underscore the significant influence of vaccination-related parameters, particularly *ϕ, η* and *θ*, on both ℛ_*c*_ and the total number of active infections. This implies that strategies aimed at enhancing vaccination outcomes, whether through augmenting vaccination rates, extending the duration of vaccine-induced immunity, or fortifying the protective efficacy of vaccines, can serve as effective measures in curtailing ℛ_*c*_ and mitigating infections. This insight provides valuable guidance for formulating and refining vaccination strategies in the battle against infectious diseases.

**Figure 4.**
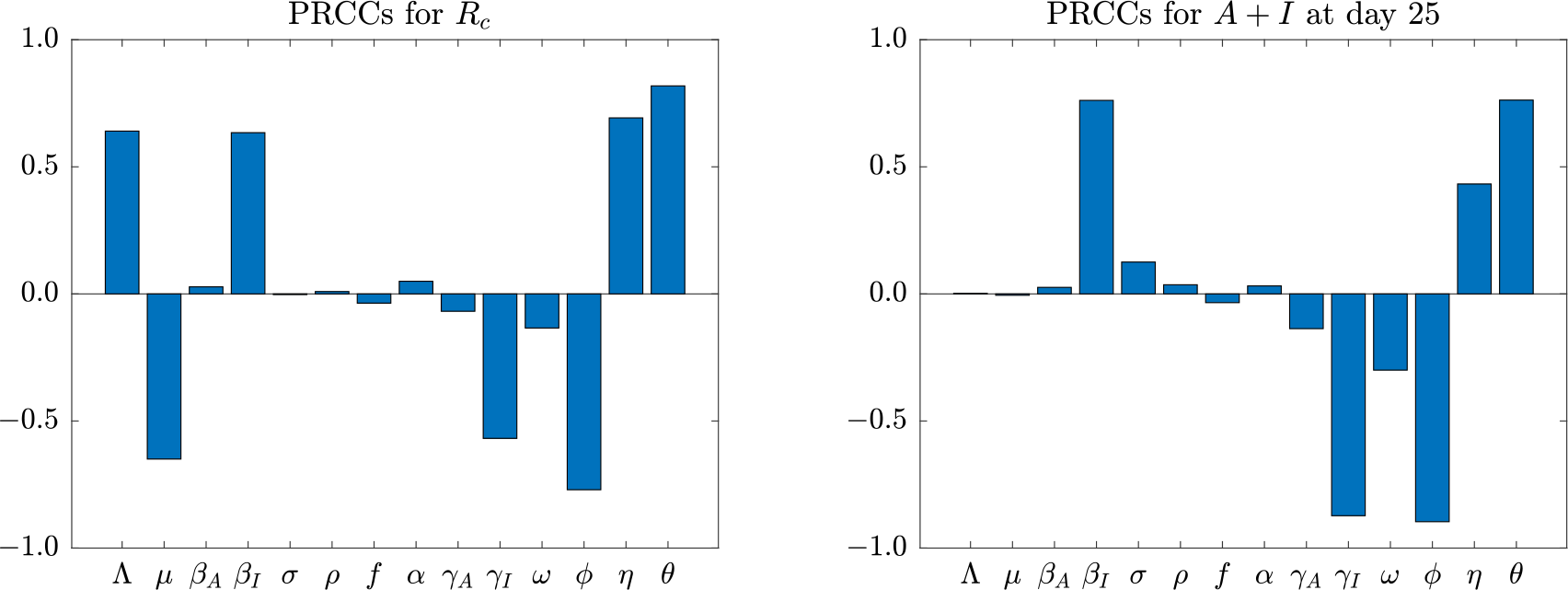
Partial Rank Correlation Coefficients (PRCCs) for the control reproduction number ℛ_c_ and active infections A + I on day 25 are assessed with respect to all parameters. The baseline values for all parameters are provided in Table 1, and for most of them, the ranges extend 20% both to the left and right of their baseline values.

Given the substantial impact that the vaccination rate *ϕ*, vaccine waning rate *η*, and vaccine escape proportion *θ* have on the disease dynamics (Figure 4), we have generated contour maps illustrating the control reproduction number ℛ_*c*_ in relation to these parameters (Figure 5). The results indicate that, under these specified parameter values, it is possible to reduce ℛ_*c*_ below the critical threshold of unity. This reduction relies on both increasing the vaccination rate and decreasing the vaccine waning rate simultaneously. However, this reduction is only achievable when *θ* assumes a small value, emphasizing the necessity of a sufficiently high level of protection provided by the vaccines. Therefore, in scenarios of virus infections, such as SARS-CoV-2 infection characterized by frequent viral mutations and limited cross-immunity, conditions which often lead to a high escape proportion, supplementary control measures may be crucial in tandem with vaccination efforts. This highlights the complexity of managing infectious diseases in contexts where the virus is prone to rapid evolution, and where standard vaccination strategies may not be adequate on their own.

**Figure 5.**
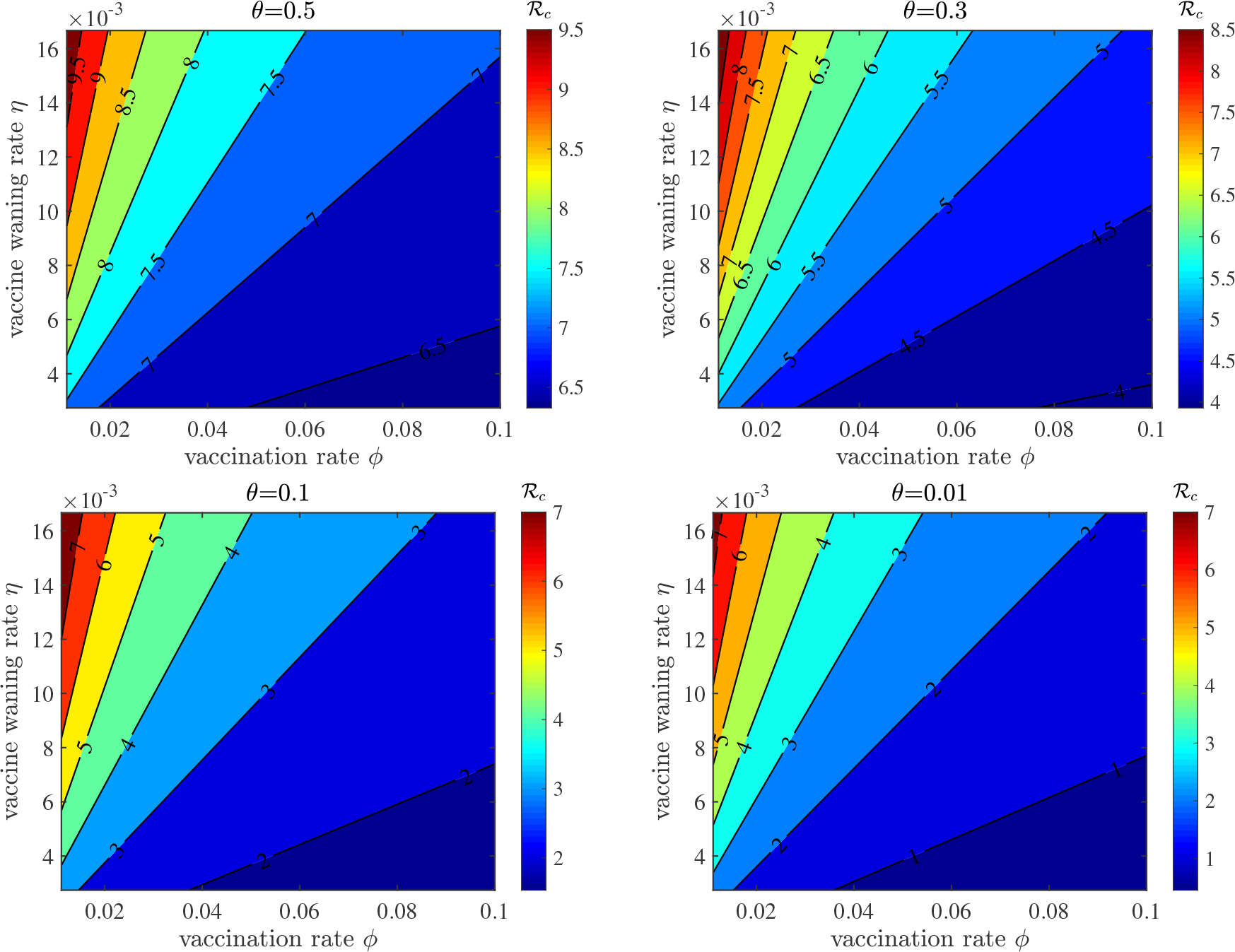
Contour maps illustrating the control reproduction number ℛ_c_ concerning the vaccination rate ϕ and vaccine waning rate η for various vaccine escape proportions θ. The remaining parameter values are provided in Table 1.

In Figure 6, we explored the projected dynamics of both asymptomatic and symptomatic infected individuals, considering varying parameters *ϕ, η*, and *θ*. The upper panels illustrate the prevalence of individuals in compartments *A* and *I* under different vaccination rates *ϕ*. All other parameters align with those outlined in Table 1. The initial condition was chosen to be the endemic equilibrium in the absence of vaccination. The results demonstrate a decrease in prevalence as vaccination rates increase in both plots. This suggests that accelerated vaccination efforts lead to a reduction in infections. The middle panels show the dynamics under varying vaccine waning rates *η*. There is a noticeable decrease in prevalence as vaccine-induced immunity endures for a longer period. Hence, an extended duration of vaccine protection results in fewer infections. In the lower panels, we presented the dynamics with different vaccine escape proportions *θ*. The findings indicate a reduction in prevalence as the immune escape parameter diminishes, which implies that higher levels of vaccine-conferred protection lead to a decrease in infections. These simulation results collectively emphasize the pivotal role of vaccination, providing invaluable insights into how adjustments in vaccination rates, the extension of vaccine-induced immunity, and the enhancement of vaccine efficacy can effectively curtail the spread of infectious diseases.

**Figure 6.**
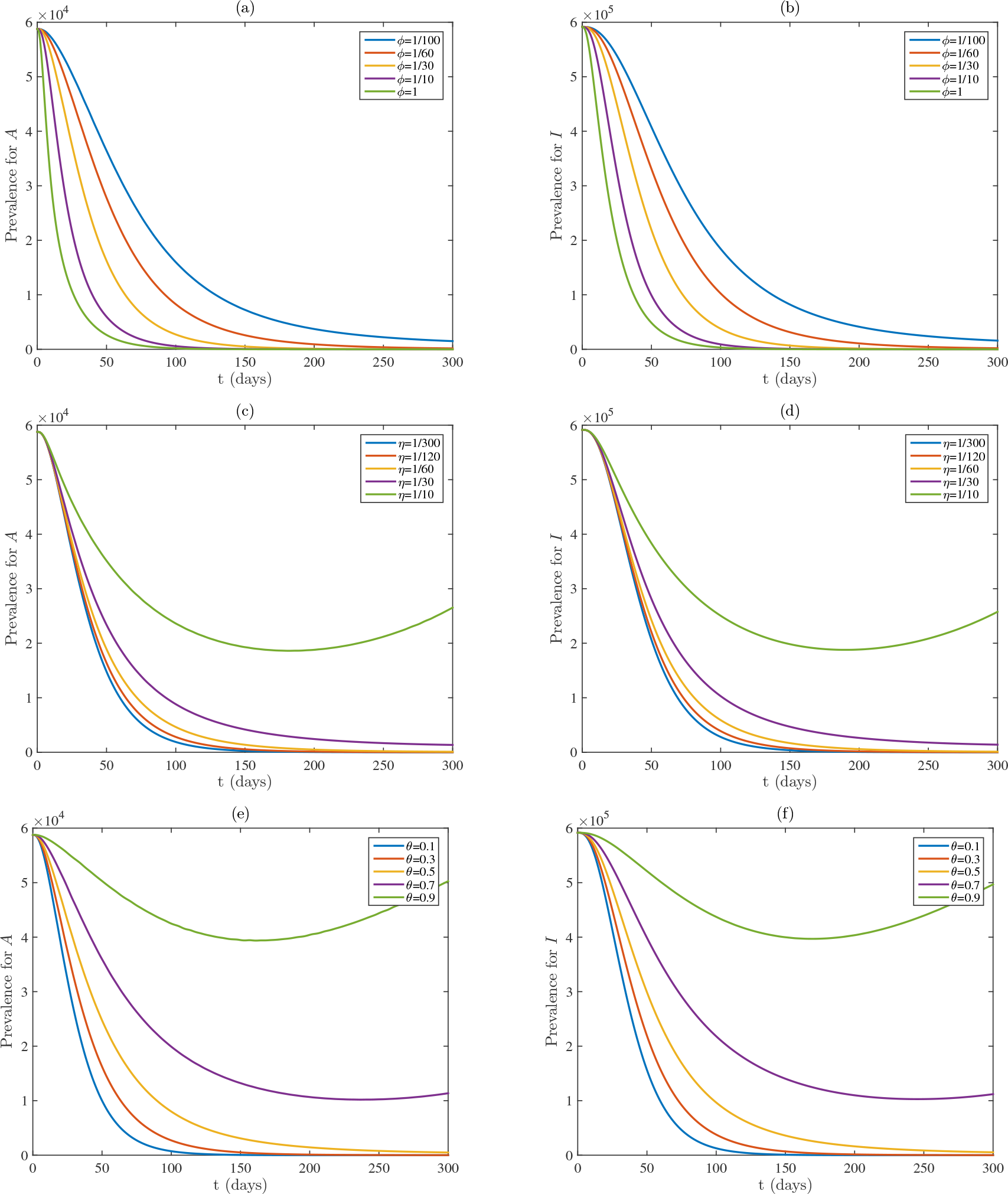
Prevalences of asymptomatic infected A and symptomatic infected I with different vaccination rates ϕ (upper panels), vaccine waning rates η (middle panels), and vaccine escape proportions θ (lower panels). Other parameter values are given in Table 1. The initial condition is the endemic equilibrium of the model without vaccination.

## 5. Conclusion and discussion

Infectious diseases present substantial challenges to the global public health. Mathematical modeling has significantly contributed to our comprehension of disease spread and the development of effective prevention and control strategies. This work advances our understanding by constructing a comprehensive mathematical model that incorporates asymptomatic transmission and waning immunity, two factors that may potentially reshape disease dynamics and impact the efficacy of interventions. Through mathematical analysis, sensitivity test and numerical investigations, we gained insights into the intricate interplay of these elements, among other factors, and their implications for disease control.

We derived the basic reproduction number ℛ_0_ for the model, which was proved to provide a critical threshold for disease extinction or persistence. When ℛ_0_ falls below 1, the disease is predicted to die out. Thus, this metric serves as a fundamental benchmark for evaluating the effectiveness of prevention and control measures. The basic reproduction number comprises contributions from both asymptomatic and symptomatic transmissions, which highlight the importance of targeting both forms of infection for effective disease control.

Numerical simulations of the model and sensitivity analysis have yielded a few results that may have profound implications for the management of infectious diseases. Firstly, we found that the level of susceptible population significantly influences disease dynamics (Figures 2 and 3). Populations with lower susceptibility tend to experience lower and delayed infection peaks. This observation underscores the importance of tailored interventions, particularly in settings where varying levels of susceptibility exist within the population. Understanding and addressing this susceptibility gradient can help optimize the allocation of resources and control measures.

Secondly, our findings indicate the pivotal role of vaccination in controlling infectious diseases. Specifically, increasing vaccination rates, extending the duration of vaccine-induced immunity, and enhancing vaccine efficacy were identified as critical strategies for reducing the control reproduction number ℛ_*c*_ below the critical threshold of unity (Figures 4, 5 and 6).

This implies that comprehensive vaccination efforts, when feasible, can play a central role in curtailing disease spread. Policymakers and health authorities should prioritize strategies that facilitate widespread vaccination, taking into account factors such as vaccine coverage, accessibility, and public perception.

However, it is essential to recognize that the effectiveness of vaccination strategies is contingent on maintaining a low vaccine escape proportion (Figure 5). In situations characterized by frequent viral mutations and limited cross-immunity, where high escape proportions are more likely, supplementary control measures may be essential alongside vaccination efforts [29]. This emphasizes the need for a multifaceted approach to disease management, especially in scenarios where the virus is prone to rapid evolution. Public health interventions should not rely solely on vaccination but should incorporate strategies to mitigate the impact of high escape proportions, such as genomic surveillance and adaptive vaccine development.

Our work has a few limitations. The mathematical model, while comprehensive, relies on a set of assumptions and simplifications. It assumes homogeneity within compartments and does not consider spatial dynamics [13, 26, 38], which can be relevant in real-world scenarios. Additionally, the model does not account for behavioral changes in response to disease outbreaks [1, 8, 14, 17], which can significantly impact transmission dynamics. Future research could explore more complex models that address these factors and their interactions.

Another limitation lies in the parameter values used in our simulations, which are based on data fitting to COVID-19 cases [21]. While this allowed us to gain some insights into the dynamics of a real-world infectious disease, it’s important to recognize that these parameters may vary in different contexts and for different pathogens. Therefore, the applicability of our findings should be considered within the specific context of the disease under investigation. Using the parameter values in Table 1, we estimated that the relative contribution to the control reproduction number from asymptomatic and symptomatic transmissions is less than 10%, which is lower than the estimates in some other studies [33, 39]. The discrepancy may be due to variations in study methodologies, population demographics, testing availability, and the definition used for asymptomatic cases. It is also worth noting that the incorporation of vaccination in the model does not alter the estimate of the relative contribution; see the formulas given in (3.3) and (3.14).

In conclusion, our modeling, analysis and numerical simulations have advanced our understanding of the intricate dynamics of infectious diseases, particularly concerning asymptomatic transmission and waning immunity. The insights gleaned from this research offer practical implications for the design and implementation of effective disease control and prevention strategies. By considering the impact of vaccination rates, the duration of vaccine-induced immunity, and vaccine efficacy, policymakers and public health officials can make informed decisions to mitigate the spread of infectious diseases and safeguard public health. Moreover, this study highlights the need for flexibility and adaptability in disease management approaches, particularly in the face of rapidly evolving pathogens. Continued research in this area, along with real-world data validation, will further refine our understanding and guide evidence-based strategies for the management of a specific infectious disease in diverse contexts.

## Data Availability

All data produced in the present work are contained in the manuscript

